# A standardized non-linear approach to studying menstrual cycle effects on brain and behavior

**DOI:** 10.64898/2026.04.10.26350619

**Authors:** Mateja Perović, Michael L. Mack

**Affiliations:** Department of Psychology, University of Toronto, Toronto, ON, Canada M5S 3G3; Centre for Addiction and Mental Health, Toronto, ON, Canada M5T 1R8

## Abstract

Menstrual cycles are major biological events with extensive effects on the brain and cognition, experienced by half of the human population. To develop a comprehensive account of human cognition, it is necessary to successfully integrate and characterize menstrual cycle effects in cognitive science research. However, current approaches to menstrual cycle analysis suffer from low data resolution and are not well-equipped to capture the highly variable, non-linear changes in outcomes of interest across the cycle. We present a validated standardized method remedying these issues, demonstrate its utility using hormonal, behavioral, and neuroimaging data, and provide an open-source toolkit to facilitate its use.

## Introduction

Half of the human population will experience menstrual cycles in their lifetime. These complex biological events have notable physiological effects beyond the reproductive system, including extensive effects on brain structure and function, and, importantly, cognition (Bernal and Paolieri, 2022; Dubol et al., 2021). Both resting state connectivity (Avila-Varela et al., 2024; Mueller et al., 2021; Pritschet et al., 2020; Taylor et al., 2020) and task-based activation (Bayer et al., 2018; Dubol et al., 2021; Pletzer et al., 2019) vary as a function of ovarian hormone changes across the menstrual cycle. Effects on hippocampal structure (Barth et al., 2016; Lisofsky et al., 2015; Protopopescu et al., 2008; Zsido et al., 2023) and function (Bayer et al., 2018; Pletzer et al., 2019; Pritschet et al., 2020) have been particularly well-documented, along with cycle-related variation in cognitive processes associated with hippocampal function, such as pattern separation (Perović et al., 2026, 2023; Perović and Mack, 2025) and visuospatial abilities (Brown et al., 2023; Hausmann et al., 2000; Peragine et al., 2020). Similarly, the menstrual cycle has been shown to modulate executive function at both behavioral (Gravelsins et al., 2021; Jacobs and D’Esposito, 2011; Louis et al., 2023) and neural (Jacobs and D’Esposito, 2011) levels.

Developing a complete account of human cognition thus necessitates deeper understanding and characterization of menstrual cycle effects, however, this critical physiological process remains neglected in cognitive neuroscience research. Less than 0.5% of the 50,000 magnetic resonance imaging papers published in the recent three decades have considered menstrual cycle effects (Jacobs, 2023a). One barrier to increased inclusion of menstrual cycle variables in research on cognition lies in methodological challenges regarding cycle definition and analysis. In fact, inconsistency in cycle measurement and analysis has been identified as a key factor underlying the mixed findings in the behavioral literature on the menstrual cycle and cognition (Bernal and Paolieri, 2022). Here, we first summarize the typical approaches and common pitfalls in menstrual cycle research, before presenting a recent advance in the form of a data-driven, broadly applicable, standardized non-linear approach with capacity for increased data-resolution and detailed characterization of cognitive and neuroimaging data across the menstrual cycle.

The typical approach to analyzing menstrual cycle effects in psychology and neuroscience involves dividing the cycle into phases based on confirmed or expected hormone levels, which are then compared to each other as discrete groups. There are several popular approaches to such division (Schmalenberger et al., 2021) (Figure 1A), each with its own strengths, however, several methodological issues emerge regardless of phase definition. Most notably, ovarian hormone levels vary across the menstrual cycle in a non-linear fashion (Figure 1A), meaning that group-based analytic approaches, even when combined with linear analyses of specific hormone effects, may inadvertently limit understanding of the underlying biological processes and overlook meaningful patterns. Given the dynamic and non-linear nature of changes in hormonal levels across the cycle, it is likely that their effects would be best characterized in a continuous, non-linear fashion.

**Figure 1.**
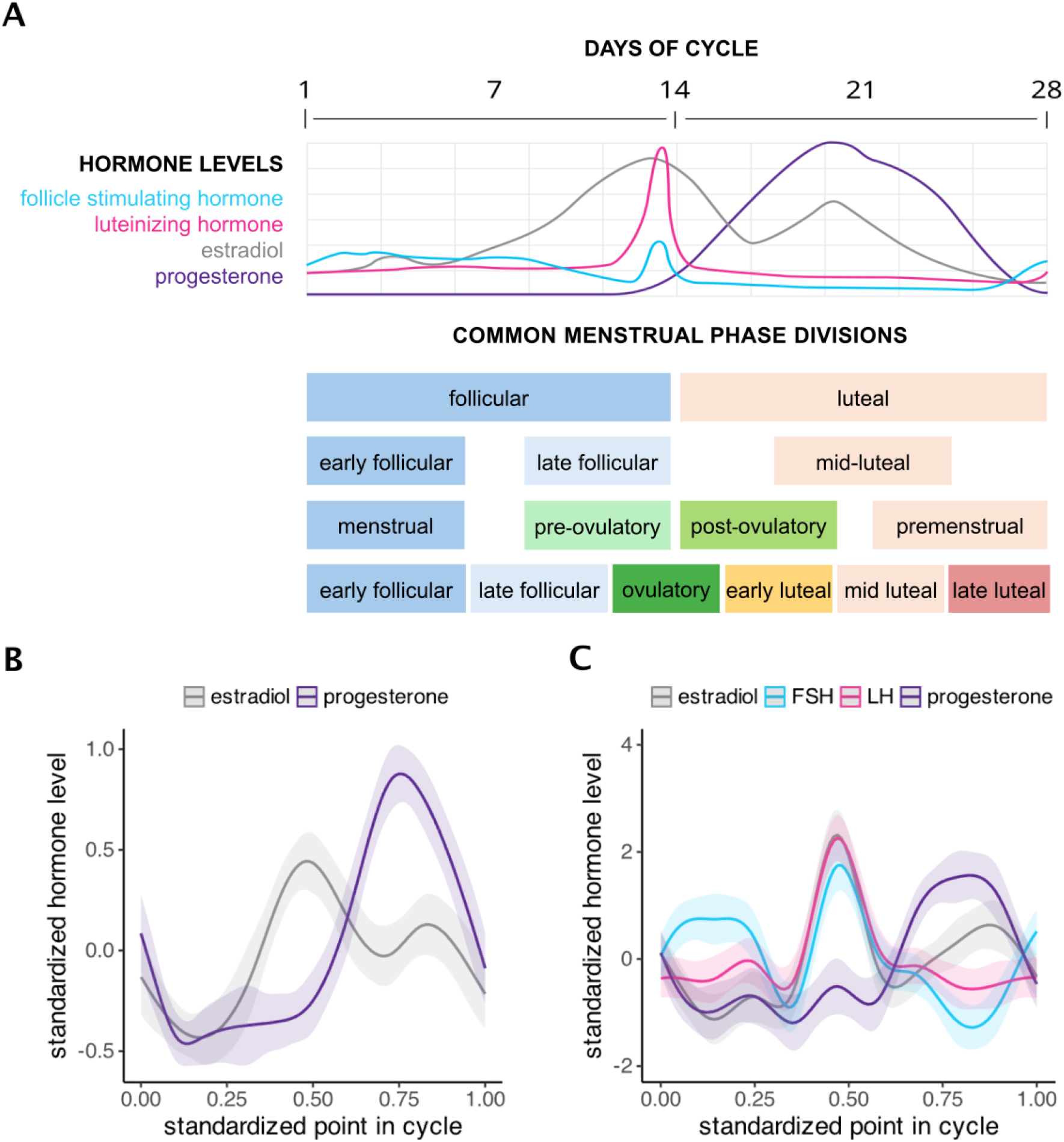
Overview of typical hormonal changes across the menstrual cycle and the corresponding variety of menstrual cycle phases commonly estimated in the menstrual cycle literature (A), along depictions of hormonal changes across the cycle as estimated by the current method in both traditional (B) and precision-phenotyping (C) cohorts, demonstrating the alignment of the current method with the non-linear nature of hormone fluctuations across the cycle. Figure depicting typical hormonal changes is adapted from (Isometrik et al., 2009).

Additionally, defining phases is not a straightforward process. Frequently, researchers use self-reported cycle information, such as length and day of cycle, to predict when participants will be in a given phase of their cycle and schedule their testing accordingly. Sometimes, ovarian hormone levels are used to confirm phase assignment. However, while hormones fluctuate across the cycle in a predictable fashion on average, there is enormous variance in cycle length and the associated hormone peaks and valleys between individuals (Gloe et al., 2023). This variance makes it challenging to recruit and test participants based on anticipated menstrual cycle phases. Furthermore, most studies do not define phases in a standardized way (Gloe et al., 2023; Schmalenberger et al., 2021) partly due to a lack of standardized normative hormone ranges for each phase (Gloe et al., 2023). One solution to these issues is to test participants multiple times and determine cycle phase based on their individual variation in hormone levels. This type of longitudinal approach is the gold standard in the literature (Gloe et al., 2023; Schmalenberger et al., 2021), however, repeated measures are not always feasible due to cost- or time-related constraints.

Finally, the phase/group-based analysis approach inevitably limits capacity for capturing within-phase variance. Even in cases where phases are carefully defined, division of the cycle into phases itself decreases statistical power by reducing the highly variable cycle phases to the level of group means and standard deviations. While this is nevertheless informative as broad judgments on differences between phases can be made, this approach fails to capture true biological variance occurring within a given phase.

As a solution to these issues, we outline a highly flexible, standardized approach to studying menstrual cycle effects on cognition (Perović et al., 2023; Perović and Mack, 2025) which treats the menstrual cycle as a scaled, continuous variable (‘cyclepoint’) with non-linear properties, mirroring the nature of hormonal fluctuations across the cycle and circumventing the need for cycle phase definition at the analysis stage. Key benefits include reduction of bias from individual variance in cycle length, increased power, and high data resolution. Critically, the current approach is entirely data-driven, making it ideal for both hypothesis testing and generation.

The toolkit accompanying this article (https://github.com/macklab/cyclepoint) provides ready-to-use code for calculation of the standardized cyclepoint, including its scaling to accommodate for generally lesser variability of the luteal phase in terms of length across cycles (Reed and Carr, 2000); analysis, including validation; and a sample pipeline for adaptation of cyclepoint to fMRI data.

This method and the accompanying toolkit are poised to help standardize menstrual cycle research on cognition, enable easier cross-lab collaboration through simplification of data harmonization pipelines, and allow for more precise characterization of outcomes of interest across the menstrual cycle. Notably, the method can be applied to both cross-sectional and repeated measures data, increasing sensitivity and statistical power in exploratory studies, and boosting data resolution of the gold-standard longitudinal approach.

### Materials and Equipment

No specialized equipment is strictly required for method implementation. In the current manuscript, hormonal, behavioral, and structural neuroimaging analyses were completed in R (version 4.0.3). Functional neuroimaging analyses were completed using R (version 4.0.3) and Python (version 3.11.4). For fMRI applications, similar considerations apply as for other types of fMRI analyses – software for preprocessing and level one analysis is required (fMRIPrep and FSL were used for the current analyses). The following two pieces of information are required for method implementation: current day of participant’s menstrual cycle, total length of participant’s menstrual cycle. We encourage researchers to ensure accuracy of these data either by asking the participant to confirm them using a period tracker app or calendar, or by longitudinally tracking participant cycle information across the full length of at least one cycle. Collection of hormone measures, including confirmation of ovulation is also highly encouraged where resources are available.

## Methods

Treatment of the menstrual cycle as a standardized, continuous variable with non-linear properties forms the basis of the current approach. To enable this, a scaled standardized point in cycle variable is calculated (cyclepoint = current day of cycle / cycle length) to be used as a main predictor variable. This results in standardization of each participant’s cycle day to a scale ranging from 0 (first day of menstrual cycle) to 1 (last day of menstrual cycle). In all analyses, effects of cyclepoint are modeled using generalized additive models (GAM) with smoothing on the cyclepoint predictor using a cyclic cubic spline basis function, and random intercepts for participants.

The GAM modeling approach has similar benefits to linear mixed modelling – which is typically recommended in gold-standard repeated-measures menstrual cycle designs (Gloe et al., 2023; Schmalenberger et al., 2021) – with the added benefit of capacity to capture non-linear effects in a data-driven fashion. A general demonstration is shown below, where Y is the outcome variable, β_0_ is the model intercept, and f_j_(x_j_) represents the smooth function of predictor x_j_. Each term f_j_(x_j_) represents the isolated contribution of that predictor.

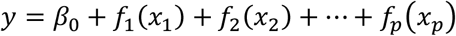

This formulation allows GAMs to flexibly capture complex relationships without overfitting. The data-driven nature of this approach makes GAM superior to other non-linear approaches such as traditional spline regression, which require researchers to inform the model of where differences in outcomes of interest might emerge. While initial iterations of the current method have utilized spline regression (Perovic et al., 2022), GAM is the preferrable analytic approach due to its flexibility and data-driven nature. Critically, GAM approaches can be used for modelling of single continuous predictors with non-linear effects, interactions between non-linear predictors and binary or categorical variables, as well as interactions between two non-linear predictors. This makes GAM highly suitable for analyzing a range of outcomes associated with the menstrual cycle. The current method utilizes a cyclic cube spline smoothing function as it has several key advantages for menstrual cycle research:

1. It tracks patterns in the data in a manner consistent with typical nonlinear patterns of hormonal variation across the menstrual cycle.
2. It respects the cyclical nature of the menstrual cycle — meaning the end of one cycle connects smoothly to the beginning of the next.
3. It avoids artificial discontinuities at the cycle boundaries (e.g., day 1 and day 28), which can otherwise introduce bias or artifacts in the model.

In the following sections, we summarize the validation process of the cyclepoint approach, starting with basic validation using hormone data across the menstrual cycle to show that the current approach is effective at capturing hormonal variations across the cycle in line with their natural, non-linear fluctuations. We then provide in-depth validation of the model for analysis of multimodal neurocognitive outcomes associated with the menstrual cycle. This includes behavioral data as well as application of the method to structural and functional neuroimaging data. Finally, we discuss further scaling options for researchers interested in tweaking the cyclepoint measure based on potential differences in cycle phase variance. For the majority of the reported analyses, the GAM model can be conceptually represented as:

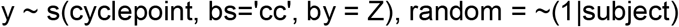

Where y is the outcome value, s(cyclepoint,bs=‘cc’) is the standardized cycle point variable, with a cyclic spline basis function, Z is the indicator for the interaction factor g (e.g., hormone, cognitive task condition, brain region of interest), and ∼(1|subject) represents the participant random effect. A key outcome of GAM analyses is EDF, or effective degrees of freedom – an indicator of non-linearity outcomes of interest. EDF of 1 indicates a linear effect and values higher than 1 indicate non-linearity.

## Results

### Basic physiological validation

To demonstrate the capacity of cyclepoint to closely track the changes in hormone levels across the menstrual cycle, we leveraged data collected in our lab as well as several published, open-access datasets (Hidalgo-Lopez et al., 2021; Pletzer and Noachtar, 2023; Taylor et al., 2020) to compile a large dataset of estradiol and progesterone measures across the menstrual cycle (N = 310, with repeated hormone measures available for n = 275). We predicted standardized hormone levels by cyclepoint using GAM with smoothing on the cyclepoint predictor with a cyclic cubic spline basis function, an interaction with a hormone factor and random intercepts for participants. Extreme hormone values (SD ± 4) were excluded from the analysis. Results (Figure 1B) indicate significant non-linear effects of cycle point on both estradiol (EDF = 5.33, *F*(8) = 38.48, *p* < .001) and progesterone levels (EDF = 5.64, *F*(8) = 96.45, *p* < .001), illustrating the capacity of the cycle point measure to track changes in hormone levels in a manner matching their typical fluctuation across the menstrual cycle (e.g., Figure 1A).

To further validate this metric, we leveraged an open-source dense-sampling dataset of a single individual(Taylor et al., 2020) in order to predict standardized ovarian hormone measures over the menstrual cycle using a GAM with a cyclic cubic spline basis function for cyclepoint. Dense-sampling has become increasingly popular within the menstrual cycle and broader women’s health literatures in recent years, as it provides opportunity for high-resolution data appropriate for tracking rapid effects of changing hormone levels across the menstrual cycle and other hormonal transitions(Jacobs, 2023b). Using this open access dataset, we provide further validation of the cyclepoint metric for granular tracking of hormonal changes (EDF_estradiol_ = 7.04, *F*(8) = 49.2, *p* < .001; EDF_FSH_ = 6.81, *F*(8) = 40.95, *p* < .001; EDF_LH_ = 7.05, *F*(8) = 37.3, *p* < .001; EDF_progesterone_ = 4.87, *F*(8) = 57.98.9, *p* < .001; Figure 1C), and demonstrate its utility for analysis of high-resolution dense-sampling data.

### Application to behavioral data

We demonstrate the benefits of the current method for behavioral analysis using the results of a previously published study (Perović et al., 2023) which demonstrates the capacity of cyclepoint to capture nuanced variation in complex cognition across the menstrual cycle, including within parts of the cycle that are typically collapsed into a single cycle phase in more traditional analyses.

In order to not distract from the methodological focus of the current manuscript, we can reframe the task as tracking performance on varying levels of item difficulty (low, moderate, high). For interested readers, the task and the results are outlined in detail in the original manuscript. Participants were recruited through the Prolific online recruiting platform and tested at a single time point. All included participants reported regular menstrual cycles, no chance of pregnancy, and no use of hormonal contraceptives or hormone replacement therapy. Most (79%) participants tracked their cycles using an app or a calendar. In the following section, we summarize our main findings, provide a bootstrap resampling validation, and indicate better fit of the proposed GAM approach relative to a more traditional linear mixed model.

Similarly to the hormone analyses, cyclepoint was calculated for each participant and its effect modeled using GAM. Relative to a baseline linear mixed-effects model with no non-linear effects on cycle point, the GAM resulted in a better fit of the data (GAM AIC: -242.6 vs. linear AIC: - 224.6). Results from the GAM demonstrated a distinctly non-linear effect on the difficult task measure of interest (EDF = 2.93, *F*(8) = 85.17, *p* < .001). Bootstrap resampling results confirmed the reliability of this effect, with 97.7% of bootstrap estimates showing > 1 EDF (EDF_boot_ = 3.86, *p* = .02, 95% CI [1.04, 6.14]). Each bootstrap iteration included resampling with one participant excluded and re-fitting the model, suggesting a robust effect of cycle point on task performance that was not driven by specific participants.

The results indicate a selective increase in performance on difficult task items across the presumed early follicular phase of the menstrual cycle, with a peak in the late follicular phase, followed by a decrease that remains stable across the presumed luteal phase (Figure 2A). These results demonstrate cyclepoint’s ability to effectively capture complex behavior and discriminate between related outcome measures across the menstrual cycle.

**Figure 2.**
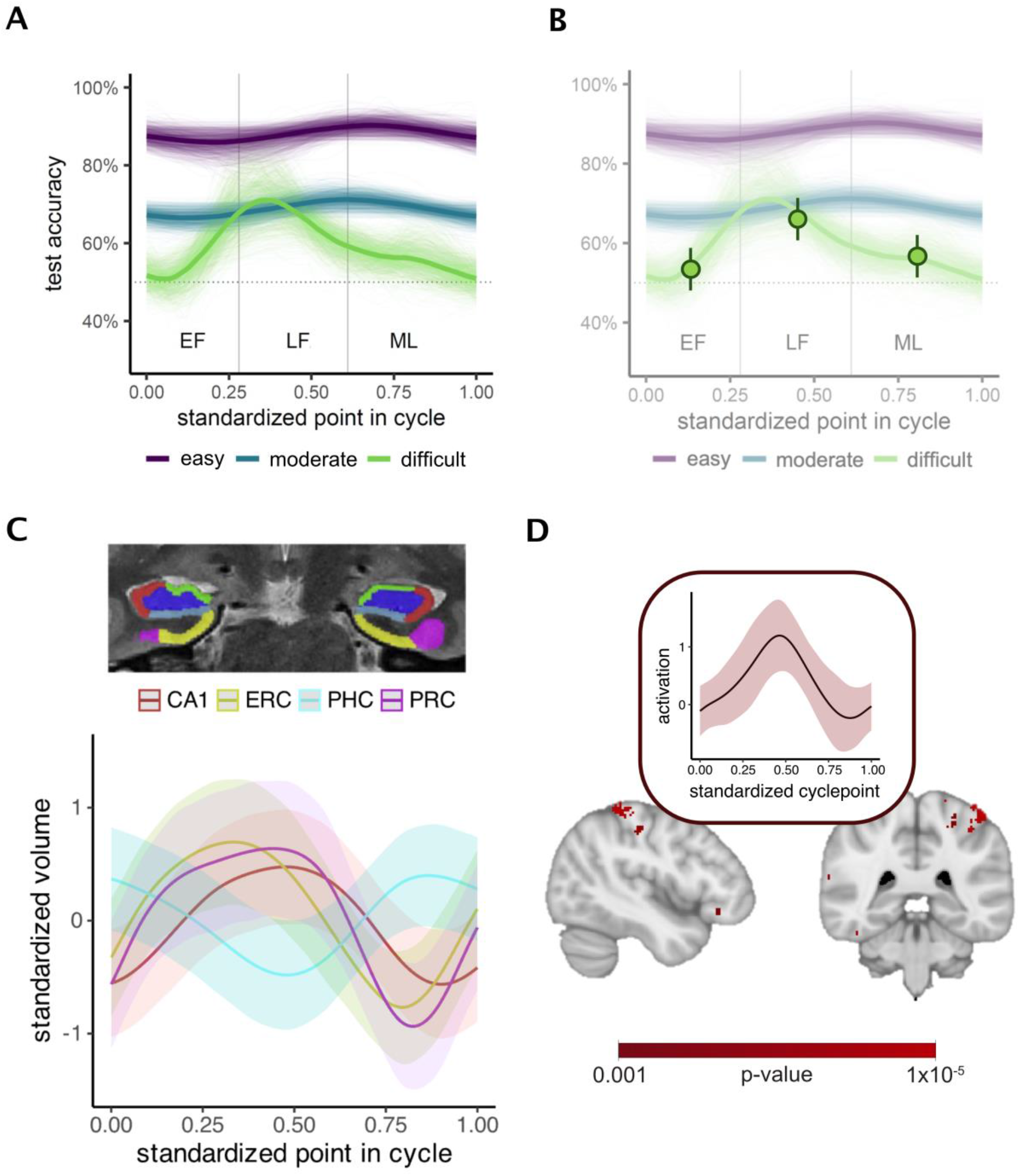
Multimodal demonstration of the current method using behavioral data indicating continuous, non-linear effects on difficult task items (in green) (A), including a visual comparison to traditional group-based analysis (B), as well as structural (C) and functional (D) magnetic resonance imaging data.

We compared these results to a traditional analytic approach in the menstrual cycle literature, whereby the cycle is divided into estimated phases (early follicular, late follicular, mid-late luteal), which are then compared to each other as discrete groups. Linear models showed findings convergent with the non-linear approach: performance was higher in the late follicular than the early follicular group (β = -0.11, SE = 0.05, t(167) = -2.31, p = .02), but it did not significantly differ from the mid-luteal group (β = -0.06, SE = 0.05, t(167) = -1.33, p = .18). The early follicular and mid-late luteal groups did not differ from each other (β = 0.05, SE = 0.05, t(167) = 0.98, p = .33). However, the nuanced changes in task performance with significant shifts occurring within phases were better captured by the GAM analysis. This benefit of GAM is illustrated in Figure 2B – while the traditional method provides similar high-level interpretation (performance is higher in the late than in the early follicular phase), cyclepoint maximizes data resolution to provide a continuous account of performance across the cycle and reveal notable within-phase dynamics (performance is not simply lower in the early follicular phase, it rises steadily until its mid-cycle peak).

### Application to neuroimaging data

Following the demonstration of the benefits of cyclepoint for characterizing behavioral data, we further validate the method for granular, continuous tracking of menstrual cycle effects in structural and functional neuroimaging research.

### Structural

Open-access data from a deep-phenotyping dataset of a single individual across a full menstrual cycle (Pritschet et al., 2020) was used for a demonstration of cyclepoint’s utility for continuous analysis of non-linear changes in hippocampal structure across the menstrual cycle. Scanning protocols are outlined in a prior publication (Taylor et al., 2020). For the current analysis, T2 scans were downloaded from the open-source repository and segmented using the Automated Segmentation of Hippocampal Subfields (ASHS) software and the Princeton Young Adult 3T Atlas for the following regions of interest within the medial temporal lobe: cornu ammonis (CA)1 and CA2/3, dentate gyrus, subiculum, entorhinal cortex, perirhinal cortex, and parahippocampal cortex (mirroring the regions of interest defined in the original analysis of the data).

Resulting volumetric measures were standardized and predicted by an interaction between the scaled cyclepoint variable and a region of interest factor, while controlling for total intercranial volume. Similar to the prior cyclepoint analyses, this analysis was modeled using a GAM with smoothing on the cyclepoint predictor using a cyclic cubic spline basis function. Results indicated non-linear effects of cyclepoint on CA1 (EDF = 1.977, F(8) = 0.98, p = .019), entorhinal (EDF = 2.33, F(8) = 1.72, p = .002), parahippocampal (EDF = 1.73, F(8) = 0.65, p = .05) and perirhinal volumes (EDF = 2.76, F(8) = 1.86, p = .002), but not on CA2/3, dentate gyrus or subiculum (all p > 0.05). This notably extended the original findings from the dataset, which showed group differences between low- and high-progesterone phases, by providing a detailed continuous account of MTL structural dynamics across the entirety of the menstrual cycle, capturing the dynamic shifts between mid-cycle and mid-luteal extrema (Figure 2C).

### Functional

Furthermore, we adapted cyclepoint to fMRI data by applying it at the level of each voxel in an exploratory whole-brain analysis screening for patterns of non-linear activation associated with the menstrual cycle. Participants (N = 41) completed MRI testing using a 3.0T Seimens Prisma Scanner, at a single point in the cycle. High-resolution T1-weighted structural volumes were acquired for co-registration. Functional images were acquired using a T2*-weighted EPI pulse sequence (TR = 2 s, TE = 2.8 ms, flip angle =7 3º, matrix = 130x130, 87 slices, 1.7 mm iso-voxels). Functional data was pre-processed using fMRIPrep, including motion correction, slice-time correction, and correction for susceptibility distortions using fieldmaps. Data were registered to participants’ T1 volume. First-level general linear model analyses were conducted in anatomical T1 space using FSL to identify voxels with unique activation to relevant task stimuli. Presentations of different stimuli types were modelled as 3s events and convolved with the double-gamma hemodynamic response function. Correct and incorrect trials (timed to occur with the onset of feedback), the 12 confound regressors, and temporal derivatives for all task regressors were included in the model. Temporal autocorrelation correction was applied using FILM pre-whitening.

We used the fist-level analyses results as inputs into exploratory GAM models which predicted voxel-level activation for relevant stimuli contrasts by cyclepoint to identify a map of regions showing non-linear, cycle-related effects. Significant clusters were defined by a voxel wise threshold of *p* = 0.001 and cluster-extent threshold of *p* = 0.05, which corresponded with a cluster extent of 29 voxels. A snapshot of the results is shown in Figure 2D, indicating a significant cluster of non-linear cycle-related activation in superior parietal and somatosensory cortices (peak MNI coordinate [-44.05, -37.75, 59.2]; EDF = 2.46, F(8) = 1.7, p = .005), a region commonly associated with the administered task. To our knowledge, this was the first application of this kind of voxel-level non-linear analysis to BOLD data – not only in the menstrual cycle literature, but within cognitive neuroscience more broadly.

### Scaling cyclepoint

The cyclepoint approach is highly robust to differences in menstrual cycle length, as it standardizes all participants to the same 0-1 cycle scale. It also circumvents the need for specific menstrual phase determination as it allows for capturing of the entirety of the menstrual cycle. However, it is still worth emphasizing that the menstrual cycle is highly variable, with evidence suggesting that the luteal phase is less variable in length than the follicular phase, generally lasting around 14 days irrespective of follicular phase length (Reed and Carr, 2000). For researchers interested in a conservative scaling approach that directly accounts for individual differences in cycle length, we provide documentation for the calculation of an adjusted cyclepoint variable.

Adjusted cyclepoint can be derived by fixing the luteal phase at 14 days and allowing the follicular phase to absorb cycle length variability. For a given total cycle length L and cycle day D, the estimated follicular phase length can be defined as F = L - 14. Cycle days occurring in the estimated follicular phase (D ≤ F) are then rescaled as cycle point_adjusted_ = (D/F) × 0.5, and cycle days in the estimated luteal phase (D > F) as cycle point_adjusted_ = 0.5 + ((D - F)/14) × 0.5. The resulting adjusted cycle point variable still standardizes each person’s cycle day to a scale ranging from 0 (first day of cycle/start of follicular phase) to 1 (last fay of cycle/end of the luteal phase), but it scales cycle mid-point (0.5) to provide a flexible adjustment for variability in cycle length and more granular estimation of the shift between follicular and luteal phases. Applied to our behavioral dataset as an example, adjusted cyclepoint performs similarly to cyclepoint (AIC_cyclepoint_= -242.6; AIC_adjusted_cyclepoint_ = -237.6), with a significant effect distinct to the difficult task items (EDF = 2.56, *F*(8) = 68.84, *p* < .001). A function for adjusted cyclepoint calculation is included in the accompanying toolkit. Where hormonal data confirming ovulation is available, the function would ideally be adjusted to scale cyclepoint based on participant-specific days of ovulation.

## Discussion

The menstrual cycle has major effects on cognition and the brain (extensively reviewed in (Bernal and Paolieri, 2022; Dubol et al., 2021; Ruehr et al., 2025)) yet research in cognitive and brain sciences rarely considers it as a predictor(Jacobs, 2023a). Here, we outline an easily applicable, scalable method for studying menstrual cycle effects on brain and behavior, and validate it using hormonal, behavioral and multimodal neuroimaging data.

The physiological validation shows that the cyclepoint approach effectively captures the dynamic and non-linear nature of hormonal changes across the menstrual cycle without a need for cycle-phase division, both in a large dataset pooled from multiple existing menstrual cycle studies as well as a precision-phenotyping dataset of a single individual. Recent work comparing a similar scaled approach to more traditional methods across 44 cycles found reduced variance in within-person hormone measures using the scaled approach (Nagpal et al., 2025). These results align with our analyses and provide further evidence of alignment between continuous approaches to menstrual cycle analysis and natural variance in hormone levels.

Beyond analysis of basic physiological data, cyclepoint provides high-resolution characterization of cognitive performance across the menstrual cycle. Published work (Perovic et al., 2022; Perović et al., 2023; Perović and Mack, 2025) demonstrates the capacity of cyclepoint to capture nuanced variation in complex cognition across the menstrual cycle, including within parts of the cycle that are typically collapsed into a single cycle phase in a more traditional analysis. Further demonstrating its sensitivity to cycle-related behavioral effects, cyclepoint effectively discriminates between related outcome measures (in the current example, task difficulty levels). Comparison of results yielded by the cyclepoint approach with the typical group-based analysis of cycle phases demonstrates advantages of cyclepoint for maximizing data resolution and capturing within-phase dynamics to provide a comprehensive account of variance in cognitive performance across the entirety of the menstrual cycle.

In line with its benefits for analysis of behavioral data, cyclepoint provides granular accounts of menstrual cycle effects on brain structure and function. We demonstrated its value for continuous tracking of structural changes in brain volume across the menstrual cycle in a deep-phenotyping sample. Additionally, we adapted cyclepoint to fMRI data by applying it at the level of each voxel in an exploratory analysis screening for patterns of non-linear activation associated with the menstrual cycle. This application demonstrates the utility of the cyclepoint approach for generating brain maps of non-linear cycle-related activation. Together, these results suggest that we can learn more about menstrual cycle effects on brain activation by treating the cycle as a continuous, non-linear variable. The cyclepoint approach is particularly valuable for neuroimaging studies, which commonly rely on smaller samples with limited statistical power. Whereas grouping participants into cycle phases further reduces power, cyclepoint enables a more sensitive assessment of menstrual cycle effects, making full use of the available data.

The current work demonstrates the capacity of cyclepoint to effectively capture the dynamic and non-linear nature of hormonal changes and associated differences in brain and behavior across the menstrual cycle. The cyclepoint approach is applicable to a broad range of data structures and sampling strategies, highly sensitive to cycle-related variance in outcomes of interest, and scalable to differences in cycle characteristics (e.g., length). Applicable to both cross-sectional and longitudinal data, the cyclepoint method increases statistical power and sensitivity in exploratory work, and maximizes data-resolution in gold standard repeated-measures studies. Application of this approach may be of particular interest to researchers studying the function of brain regions that are rich with ovarian hormone receptors and sensitive to hormonal changes across the menstrual cycle, such as the hippocampus and the prefrontal cortex (Ruehr et al., 2025). In addition to a methodological validation, the current work further highlights the importance of studying the menstrual cycle in cognitive sciences by demonstrating dynamic menstrual cycle effects on behavior, brain structure, and function.

## Data Availability

Data and code related to the manuscript are provided in the accompanying toolkit, linked in-text.

## Acknowledgements

This research was supported by NSERC Discovery Grant to MLM (RGPIN-2017-06753; RGPIN-2024-05884), Canada Foundation for Innovation and Ontario Research Fund (36601) to MLM, and Brain Canada Future Leaders in Canadian Brain Research Grant to MLM. Data and code related to the manuscript are provided in the accompanying toolkit, linked in-text.

